# Effectiveness of stay-in-place-orders during COVID-19 pandemic: Evidence from US border counties

**DOI:** 10.1101/2020.06.08.20125419

**Authors:** Srikant Devaraj, Pankaj C. Patel

**Affiliations:** Center for Business and Economic Research, Miller College of Business, Ball State University, 2000 W. University Ave., Muncie, IN 47306, USA.; Villanova School of Business, Villanova University, 800 E. Lancaster Avenue, Villanova, PA 19085, USA

**Keywords:** COVID-19, Stay-in-place-orders, border counties

## Abstract

Recent studies on US counties, with varying effect sizes, show that stay-in-place-orders (SIPOs) are associated with a decline in new COVID-19 cases. Our estimation approach relies on county-pairs across state-borders where one state has SIPO whereas the other state does not, controls for matched county-pair fixed effects and day of observation fixed-effects. The county-pair sample from southern, mid-western, and mountain region states (from March 1, 2020 to April 25, 2020) shows that daily COVID-19 incidence case growth rate is 1.994 percentage points lower for counties in SIPO states relative to those bordering in non-SIPO states. Specifically, we find SIPO reduced daily growth rates by 1.97, 2.14, 2.03, and 2.27 percentage points after 1 to 5 days, 6 to 10 days, 11 to 15 days, and 16 to 20 days, respectively. Our effect sizes are much smaller than in the previous studies with the caveat that states in the northeast and on the west coast could not be included in the border county-pair specification. We find limited evidence of heterogeneous effects in counties with a higher population density, percentage of black or Hispanic residents, proportion of population over 65 years, and social association rates in a county. Nor do we find evidence of meaningful differences in effects of SIPO by county Gini index, unemployment, or GDP. The results of this study could further inform policymakers in making decisions on SIPO extensions or lifting of such orders.

**JEL:** H75; I18

## 1. INTRODUCTION

As of May 29, 2020, the US had 1.77 million confirmed COVID-19 cases and 104,000 deaths. Though school closures and proactive work-from-home policies were some of the early responses, states across the US, to varying degrees, have implemented stay-in-place orders (SIPO). SIPO is espoused to lower daily average number of secondary COVID-19 cases per infected case. On the one hand, according to one estimate, if no preventative measures were taken about 80 percent of Americans would be infected by COVID-19 (Kissler, Tedijanto, Lipsitch, & Grad, 2020). SIPO aims to increase social distancing, an approach supported in simulation models (Teslya et al., 2020) and during past epidemics (Bootsma & Ferguson, 2007; Chinazzi et al., 2020; Ferguson, Laydon, & Nedjati Gilani, 2020). On the other hand, the economic cost of SIPO is staggering. As of May 28, 2020 40 million Americans have filed for unemployment claims, and the US GDP contracted by 5.0% in the first quarter of 2020. Increasingly there are calls to balance the economic, physiological, and psychological toll of SIPO against its impact on the decline in the spread of COVID-19.

To facilitate cost-benefit analysis of SIPO for policymakers, recent studies have provided state- and county-level estimates of SIPO on COVID-19 spread. Using differences-in-differences specification for the state-level mobility measures, Abouk and Heydari (2020) find that SIPO had the strongest impact on lowering social interactions, and in turn, it led to a steady decline in cases reaching 37% after fifteen days of implementation. Gupta et al. (2020) found that social distancing was not driven mainly by government mandates, but by early actions such as the announcements of school closures led to the increased time spent at home from 9.1 to 13.9 hours. Dave, Friedson, Matsuzawa, and Sabia (2020), using daily state-level coronavirus case data found that cumulative COVID-19 cases fell by 44 percent approximately three weeks following the adoption of a SIPO. Using cross-sectional state-level data Orazem (2020) and Reilly (2020) do not find evidence that SIPO inhibited COVID-19 case growth. Siedner et al. (2020) using interrupted time-series on state-level data find that the mean daily COVID-19 growth rate decreased by an additional 0.8% per day starting four days after the implementation of state SIPO. Rojas et al. (2020), using March 15-21 and March 22-28 unemployment insurance claims infer that labor market slowdowns were driven by school closures as a response to evolving COVID-19 conditions and state mitigation policies had a small effect.

Focusing on the densely populated counties in Texas, Dave, Friedson, Matsuzawa, Sabia, and Safford (2020) find a decline of 19 to 26 percentage in COVID-19 case growth two-and-a-half weeks following adoption. Using a synthetic control method, Friedson, McNichols, Sabia, and Dave (2020) find that one month following California’s statewide SIPO, COVID-19 cases declined by 125.5 to 219.7 per 100,000 population. Courtemanche, Garuccio, Le, Pinkston, and Yelowitz (2020) in a sample of 3,138 counties over 58 days from March 1, 2020, to April 27, 2020, found social distancing measures lowered daily growth rate by 5.4, 6.8, 8.2, and 9.1 percentage points after 1–5 days, 6–10 days, 11–15 days, and 16–20 days, respectively. In a sample of 8 Iowa counties bordering 7 Illinois counties, before and after Illinois issued SIPO on March 21, 2020, the differences in cases were −0.51, −1.15, and −4.71 per 10,000 residents after 10, 20, and 30 days, respectively. Overall, studies to date have focused on state, county, and high population density counties and the effect sizes vary significantly across studies.

We propose a cross-border county pair design for states with and without SIPO. The proposed design allows for additional advantages by controlling for spatial heterogeneity across border counties. We add matched-pair county fixed-effects to control for potential spillover conditions across state borders between neighboring counties. We control for the day-of-observation fixed-effects to account for the daily effects of COVID-19 in a fluid situation where daily national and local press-conferences may lead to changes in voluntary isolation behaviors. We further estimate standard errors by three-way clustering at state-border × county-pair × day.

Our daily growth rate estimates are much smaller than those found in prior county-level studies. We find that the effect is about 1.994 percentage points lower for counties in SIPO states relative to those in non-SIPO states. This study further adds to the ongoing evidence on SIPO and COVID-19 decline rates to further improve the generalizability of the estimates (Eichenbaum, Rebelo, & Trabandt, 2020). Upward biased estimates could lead to significant diffidence among policymakers and downward biased estimates could increase overconfidence and premature opening of the economy. Our findings do not run counter to prior studies finding support for decline COVID-19 infection rates, however, our findings show much smaller effect sizes in daily incidence growth rates and therefore call for a closer assessment of the degree of effect of SIPO. In an additional analysis, we find negligible differences in daily incidence growth rates for SIPO by county GDP, income inequality, unemployment, population density, percent black population, percent Hispanic population, or percent elderly.

How soon SIPO should be relaxed or lifted has economic and epidemiological implications. With soaring economic costs of lockdown, rising unemployment, and shrinking GDP estimates, on the one hand, there are increasing calls for opening the economy at a faster pace. On the other hand, opening the economy too soon could have a much higher human toll. Identifying the effect of SIPO on a decline in COVID-19 cases is an important piece of the cost-benefit calculus for opening the economy. Our study adds to the recent body of evidence on the extent of reduction in COVID-19 spread in the US.

## 2. METHODS

### 2.1 Data

We obtain daily data on COVID incidence at the county-level from USAFacts. USAFacts collates data from the Center for Disease Control and Prevention (CDC), state and local public health agencies.^1^ We collect dates when SIPO was effective for each state from the New York Times COVID-19 portal.^2^ All county-level economic, demographic and health variables were obtained from County Health Rankings, Bureau of Economic Analysis, and the American Community Survey.

We first create a daily balanced panel of counties between Mach 1, 2020, and April 25, 2020. We chose the end date of our analysis as April 25, 2020, because some states started partial reopening on April 26, 2020.^3^ Our treated group consists of counties from states that implemented SIPO and the control group consists of counties from the non-SIPO state that borders with at least one treated state. Based on the National Bureau of Economic Research’s neighbor county pair data, we first drop all counties that do not share a border with adjacent states. We then pair each border county of SIPO state with the closest county from the non-SIPO border state by centroid distance. After applying these filters, we had 143 counties in the treated state and 135 counties in control state. We were able to obtain 182 county pairs from these 278 border counties and 15,568 county-day observations for our analysis.^4^ Figure 1 shows the SIPO and non-SIPO counties used in our analysis and Appendix Table A lists the states used in the analysis.

**Figure 1.**
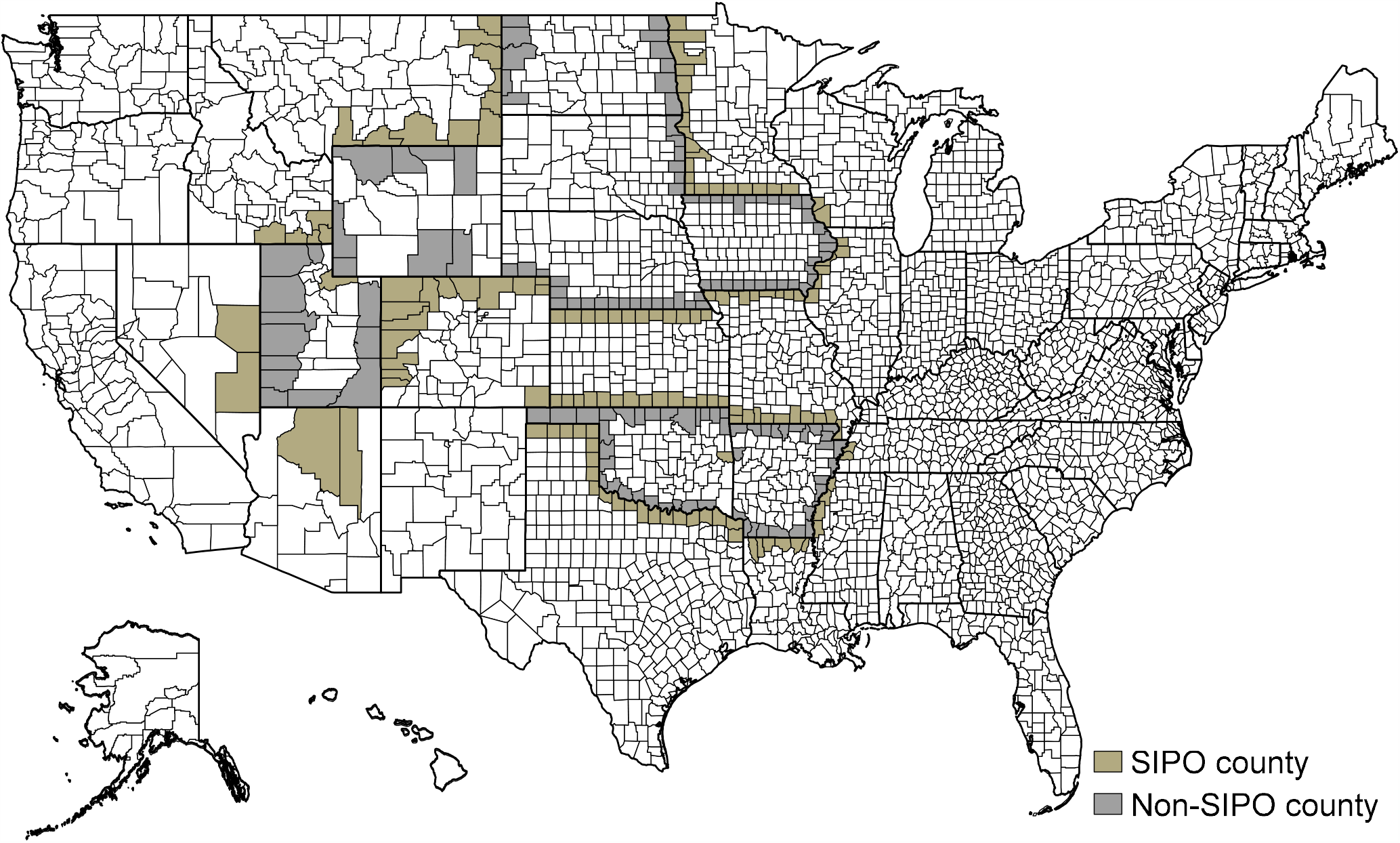
Border county - Treated and Control group. **Treated (SIPO) states** include Arizona, Colorado, Idaho, Illinois, Kansas, Louisiana, Minnesota, Missouri, Mississippi, Montana, Nevada, Tennessee, Texas, Wisconsin. We also include Sequoyah county (Oklahoma) and Summit County(Utah) in treated group as they implemented SIPO **Control (non-SIPO) states** include Arkansas, Iowa, North Dakota, Nebraska, Oklahoma (rest of border counties), South Dakota, Utah (rest of border counties), Wyoming.

### 2.2 Empirical Specification

Our identification strategy relies on border county pair analysis that controls for spatial heterogeneity between the county pairs. We follow Dube, Lester, and Reich (2010) and Peng, Guo, and Meyerhoefer (2019) and use the following specification:

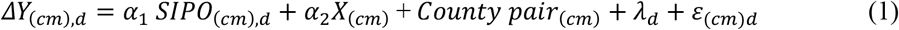

Where *c* is the county, *m* is the adjacent county across the state border, and *d* is the day. Following Courtemanche et al. (2020), our outcome variable, *ΔY*, represents change in natural log of COVID-19 cumulative incidence cases in a county between day *d* (current day) and previous (day *d-1*).

The coefficient, α_1_, is the effect of SIPO implementation on changes in COVID-19 incidences relative to the adjacent county in the pair. *X*_(cm),t_ is a vector of county-level economic, demographic, and health related control variables. We include the log of county GDP in 2018, the unemployment rate in 2018, the log of median household income in 2018, and Gini inequality index in 2018. We include 2018 county-level share of 65 years and older population, the share of African Americans in the county population, the share of the Hispanics in the county population, and the share of females in the population. We also include the share of the county population who lived in rural areas in 2010 (latest available year), social association rates in 2017, and share of the uninsured population in 2017. To control for the overall health in the county, we include a share of the population with poor or fair health in 2017, share of smokers in 2017, and share of the population who are obese in 2016.

We also include county-pair fixed effects (*Country pair*_(cm),t_) and day fixed effects (λ_d_). We use a three-way clustering of our standard errors by county-pair × state × day to control for a shared variance by neighboring counties. As a robustness test we also control for two-way clustering of standard errors by county-pair × day and by state × day. We weight all the models by county population from 2019 US census estimates. The descriptive statistics of all variables (and sources) are shown in Table 1 for both the treated and control group.

**Table 1.** Descriptive statistics

| Variables | Data source | Non-SIPO state<br>(N=7,560 county days) |  | SIPO state<br>(N=8,008 county days) |  |
| --- | --- | --- | --- | --- | --- |
|  |  | Mean | Std. Dev. | Mean | Std. Dev. |
| Daily COVID-19 exponential incidence growth rate | USAFacts | 3.3554 | 12.7239 | 3.5858 | 13.1542 |
| Log of Real GDP in 2018 | Bureau of Economic Analysis | 13.3499 | 1.1248 | 13.2146 | 1.1585 |
| Unemployment rate in 2018 | County Health Rankings / Bureau of Labor Statistics | 3.2611 | 1.0485 | 3.6404 | 1.3727 |
| Log of Median Household Income in 2018 | County Health Rankings / Small Area Income and Poverty Estimates | 10.8222 | 0.1888 | 10.8064 | 0.1994 |
| GINI inequality Index 2018 | American Community Survey | 0.4377 | 0.0353 | 0.4385 | 0.0331 |
| Share of population age 65 and over in 2018 | County Health Rankings / Census population estimates | 19.7745 | 4.4273 | 20.0742 | 4.4175 |
| Share of Black/African American population in 2018 | County Health Rankings / Census population estimates | 5.1087 | 11.7445 | 5.6495 | 13.9133 |
| Share of the Hispanic population in 2018 | County Health Rankings / Census population estimates | 6.9834 | 6.6186 | 9.8735 | 11.4788 |
| Share of female population in 2018 | County Health Rankings / Census population estimates | 49.7384 | 1.6119 | 49.6649 | 1.8486 |
| Share of population in rural areas in 2010 | County Health Rankings / Census population estimates | 64.6238 | 29.6709 | 69.2361 | 28.8301 |
| Social association rate in 2017 | County Health Rankings / Number of membership associations per capita from county business patterns | 13.6083 | 7.3380 | 15.3052 | 8.9737 |
| Share of uninsured in 2017 | County Health Rankings / Small Area Health Insurance Estimates | 11.5696 | 5.0811 | 13.1217 | 5.7985 |
| Percentage fair or poor health in 2017 | County Health Rankings / CDC-BRFSS | 16.9711 | 4.8152 | 17.2410 | 4.7445 |
| Share of smokers among the population in 2017 | County Health Rankings / CDC-BRFSS | 16.7252 | 3.2169 | 16.9307 | 3.5984 |
| Share of adults who are obese in 2016 | County Health Rankings / CDC-BRFSS | 33.9037 | 4.6683 | 32.0881 | 5.8326 |

## 3. RESULTS

### 3.1 Main results

Table 2 presents the results of the impact of SIPO on the daily COVID-19 incidence growth rate. Model 1 is the baseline model without controls but includes county-pair and day fixed-effects and three-way clustering of standard errors. Model 2 adds regional economic controls to the baseline model. Models 3 and 4 add regional demographic and health controls, respectively. Models 5 and 6 show the estimates with two-way clustering of standard errors by county-pair × day and state × day, respectively. Finally, as a robustness test, Model 7 presents the results with county fixed effects, instead of county-pair fixed effects.

**Table 2.**
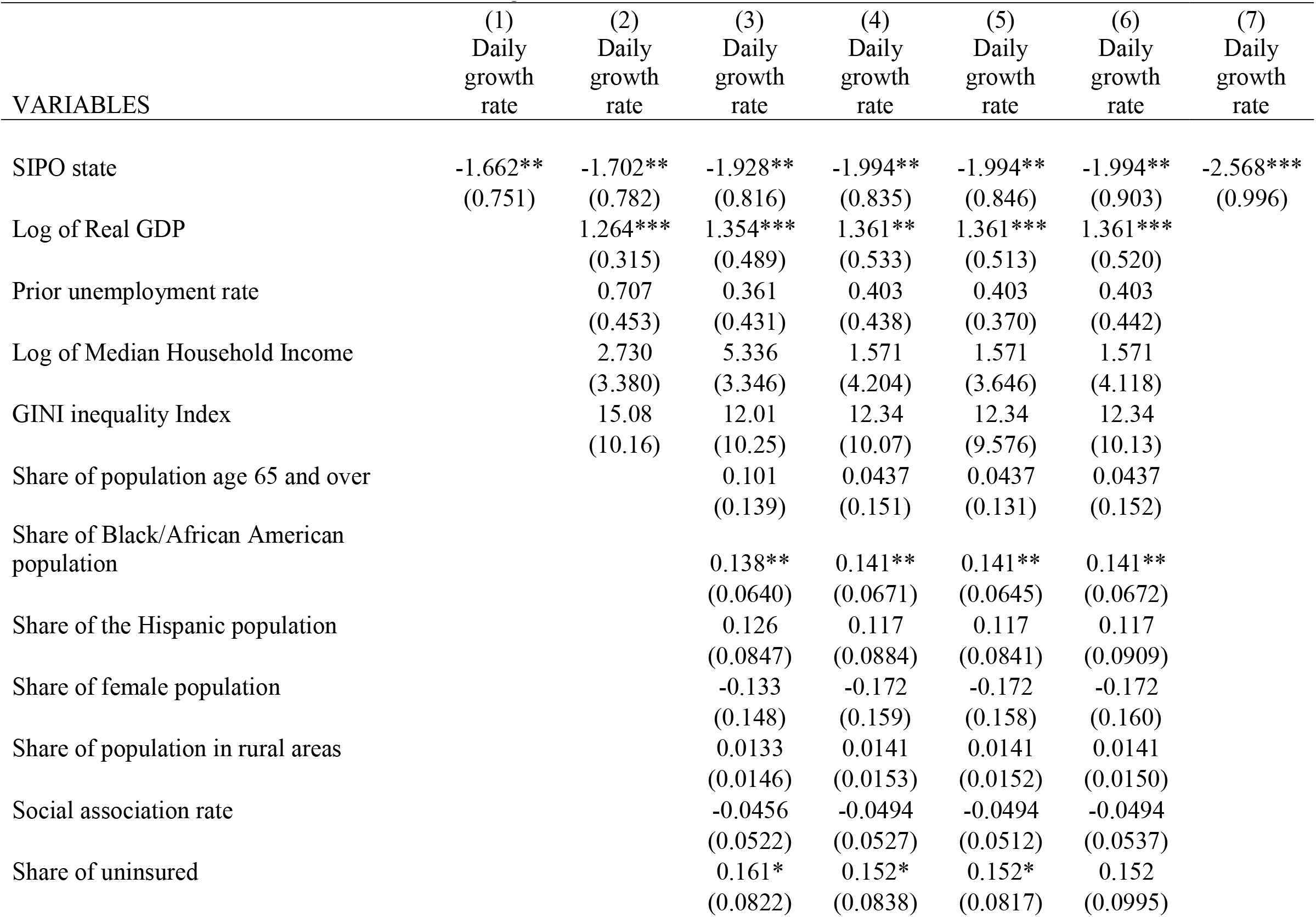
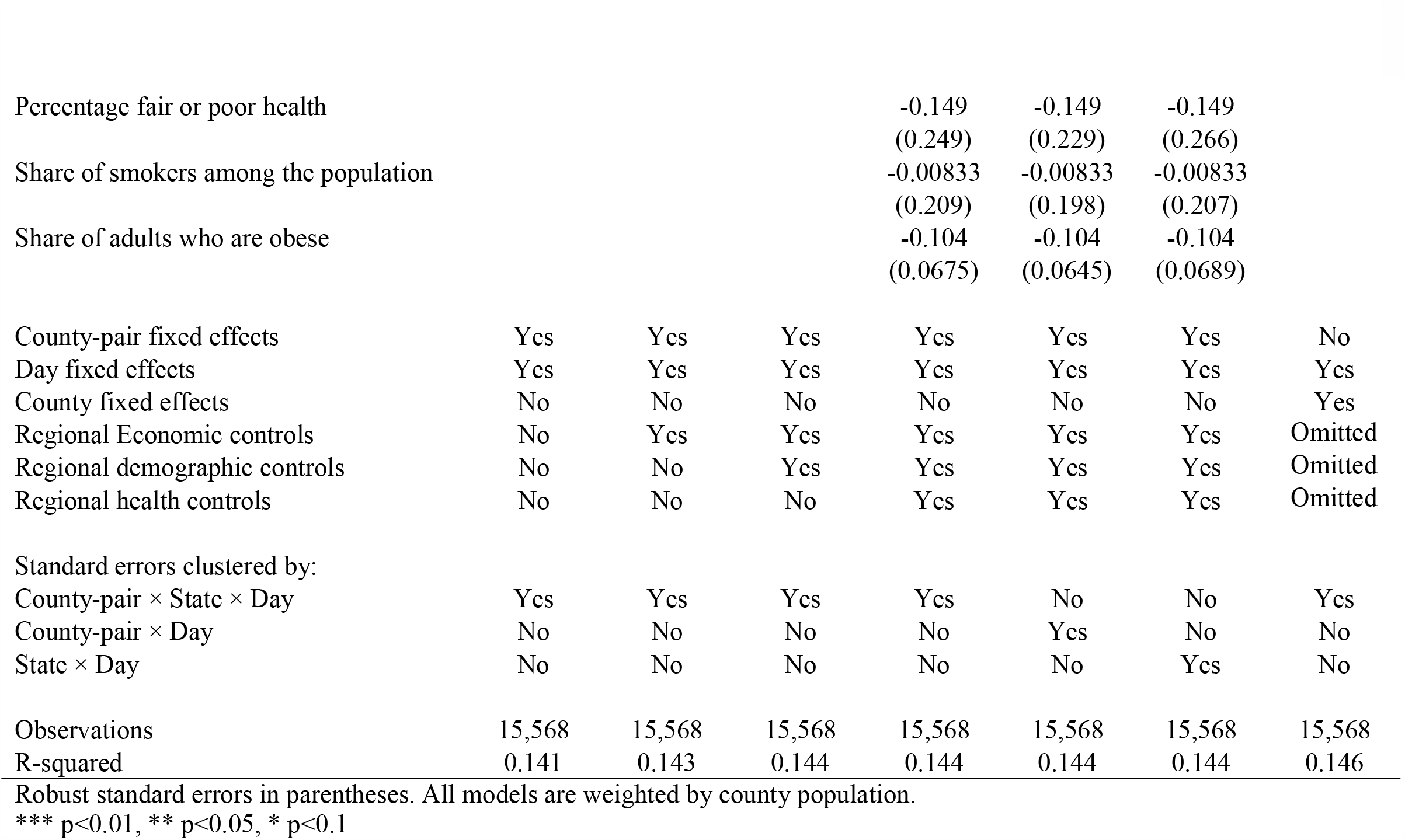
Effects of SIPO on COVID-19 incidence growth rate

| VARIABLES | (1)<br>Daily<br>growth<br>rate | (2)<br>Daily<br>growth<br>rate | (3)<br>Daily<br>growth<br>rate | (4)<br>Daily<br>growth<br>rate | (5)<br>Daily<br>growth<br>rate | (6)<br>Daily<br>growth<br>rate | (7)<br>Daily<br>growth<br>rate |
| --- | --- | --- | --- | --- | --- | --- | --- |
| SIPO state | -1.662**<br>(0.751) | -1.702**<br>(0.782) | -1.928**<br>(0.816) | -1.994**<br>(0.835) | -1.994**<br>(0.846) | -1.994**<br>(0.903) | -2.568***<br>(0.996) |
| Log of Real GDP |  | 1.264***<br>(0.315) | 1.354***<br>(0.489) | 1.361**<br>(0.533) | 1.361***<br>(0.513) | 1.361***<br>(0.520) |  |
| Prior unemployment rate |  | 0.707<br>(0.453) | 0.361<br>(0.431) | 0.403<br>(0.438) | 0.403<br>(0.370) | 0.403<br>(0.442) |  |
| Log of Median Household Income |  | 2.730<br>(3.380) | 5.336<br>(3.346) | 1.571<br>(4.204) | 1.571<br>(3.646) | 1.571<br>(4.118) |  |
| GINI inequality Index |  | 15.08<br>(10.16) | 12.01<br>(10.25) | 12.34<br>(10.07) | 12.34<br>(9.576) | 12.34<br>(10.13) |  |
| Share of population age 65 and over |  |  | 0.101<br>(0.139) | 0.0437<br>(0.151) | 0.0437<br>(0.131) | 0.0437<br>(0.152) |  |
| Share of Black/African American population |  |  | 0.138**<br>(0.0640) | 0.141**<br>(0.0671) | 0.141**<br>(0.0645) | 0.141**<br>(0.0672) |  |
| Share of the Hispanic population |  |  | 0.126<br>(0.0847) | 0.117<br>(0.0884) | 0.117<br>(0.0841) | 0.117<br>(0.0909) |  |
| Share of female population |  |  | -0.133<br>(0.148) | -0.172<br>(0.159) | -0.172<br>(0.158) | -0.172<br>(0.160) |  |
| Share of population in rural areas |  |  | 0.0133<br>(0.0146) | 0.0141<br>(0.0153) | 0.0141<br>(0.0152) | 0.0141<br>(0.0150) |  |
| Social association rate |  |  | -0.0456<br>(0.0522) | -0.0494<br>(0.0527) | -0.0494<br>(0.0512) | -0.0494<br>(0.0537) |  |
| Share of uninsured |  |  | 0.161*<br>(0.0822) | 0.152*<br>(0.0838) | 0.152*<br>(0.0817) | 0.152<br>(0.0995) |  |
| Percentage fair or poor health |  |  |  | -0.149<br>(0.249) | -0.149<br>(0.229) | -0.149<br>(0.266) |  |
| Share of smokers among the population |  |  |  | -0.00833<br>(0.209) | -0.00833<br>(0.198) | -0.00833<br>(0.207) |  |
| Share of adults who are obese |  |  |  | -0.104<br>(0.0675) | -0.104<br>(0.0645) | -0.104<br>(0.0689) |  |
| County-pair fixed effects | Yes | Yes | Yes | Yes | Yes | Yes | No |
| Day fixed effects | Yes | Yes | Yes | Yes | Yes | Yes | Yes |
| County fixed effects | No | No | No | No | No | No | Yes |
| Regional Economic controls | No | Yes | Yes | Yes | Yes | Yes | Omitted |
| Regional demographic controls | No | No | Yes | Yes | Yes | Yes | Omitted |
| Regional health controls | No | No | No | Yes | Yes | Yes | Omitted |
| Standard errors clustered by: |  |  |  |  |  |  |  |
| County-pair × State × Day | Yes | Yes | Yes | Yes | No | No | Yes |
| County-pair × Day | No | No | No | No | Yes | No | No |
| State × Day | No | No | No | No | No | Yes | No |
| Observations | 15,568 | 15,568 | 15,568 | 15,568 | 15,568 | 15,568 | 15,568 |
| R-squared | 0.141 | 0.143 | 0.144 | 0.144 | 0.144 | 0.144 | 0.146 |

Robust standard errors in parentheses. All models are weighted by county population.
\*\*\* $p < 0.01$ , \*\* $p < 0.05$ , \* $p < 0.1$

We find that SIPO significantly reduced the daily growth rate of COVID-19 incidences relative to the cross-border county in the non-SIPO state. From our preferred specification (Model 5), we find an overall reduction in daily growth rate to be 1.99 percentage points in county with SIPO state relative to the county in non-SIPO state. We find that the results are consistent across alternate standard error estimates.

### 3.2 Effects by different cut-offs post-SIPO

We also test whether there is a differential effect of SIPO on daily growth rates after a certain cut-off period. We follow Courtemanche et al. (2020) to identify specific cut-offs post SIPO for our analysis such as 1 to 5 days; 6 to 10 days; 11 to 15 days; 16 to 20 days; and 21 or more days after SIPO.

Our model specification for the cut-offs is as below:

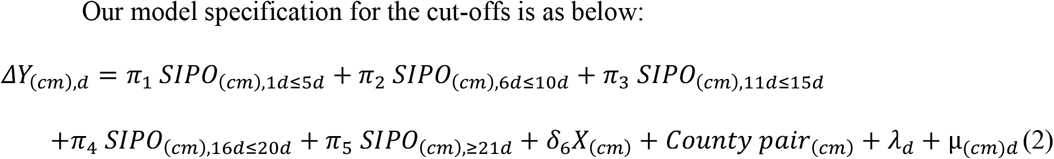

Where *c* is the county, *m* is the adjacent county across the state border, and *d* is the day.

Table 3 presents the results of our analysis. We find that the effects of SIPO on daily incidence growth rates are statistically significant for 6 to 10 days after implementation and beyond in a range of 1.97 to 2.27 percentage point decline.

**Table 3.** Effects by various cut-offs post-SIPO

| VARIABLES | (1)<br>Daily growth rate |
| --- | --- |
| 1-4 days after SIPO | -1.247<br>(1.220) |
| 6-10 days after SIPO | -1.967**<br>(0.988) |
| 11-15 days after SIPO | -2.142**<br>(0.983) |
| 16-20 days after SIPO | -2.027**<br>(0.949) |
| 21 or more days after SIPO | -2.274**<br>(1.093) |
| Observations | 15,568 |
| R-squared | 0.144 |

Robust standard errors clustered by County-pair × State × Day in parentheses. All economic, demographic, and health controls are included. Models are weighted by county population.
\*\*\* $p < 0.01$ , \*\* $p < 0.05$ , \* $p < 0.1$

### 3.3 Event study

Using two event study specifications with pre-SIPO and post-SIPO implementation cut-offs, we test whether the pre-existing trends in daily cumulative incidence rates before SIPO implementation are present relative to the adjacent county pair.

Our first event study specification includes a dummy variable for 13 or more days before the SIPO with the reference category being 1 to 12 days before SIPO:

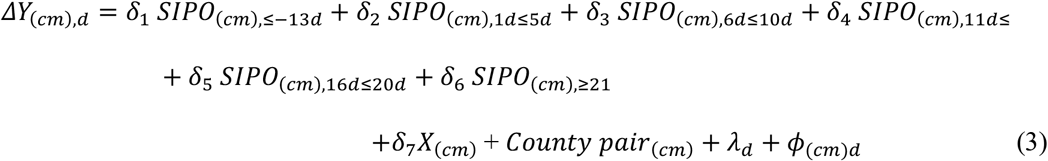

Where *c* is the county, *m* is the adjacent county across the state border, and *d* is the day.

Our second model of event study uses two dummy variables 6 to 9 days before the SIPO and 10 or more days before the SIPO:

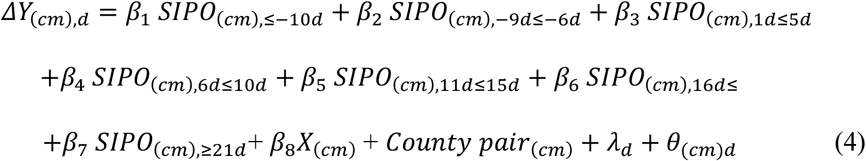

Where *c* is the county, *m* is the adjacent county across the state border, and *d* is the day.

Table 4 presents the results of the event study models. Model 1 corresponds to results from equation (3) and Model 2 corresponds to results from equation (4). Figure 2 shows the event study plots for both these models. We find that the coefficients are insignificant before the SIPO implementation and during 1 to 5 days post-SIPO relative to the reference category (1 to 12 days before SIPO). We find significant effects on daily growth rates from 6 to 10 days post-SIPO and beyond.

**Table 4.**
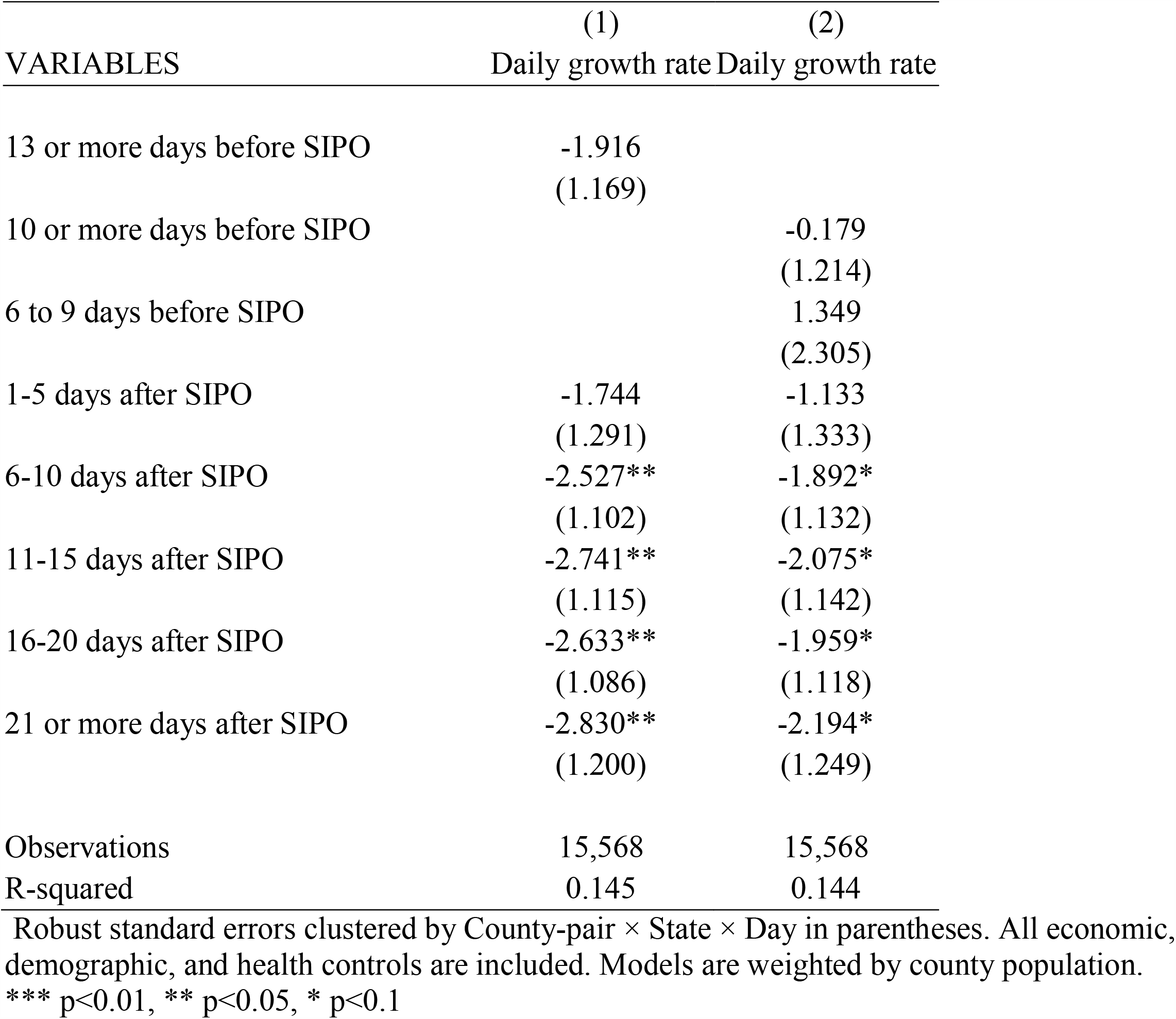
Event study estimates

| VARIABLES | (1)<br>Daily growth rate | (2)<br>Daily growth rate |
| --- | --- | --- |
| 13 or more days before SIPO | -1.916<br>(1.169) |  |
| 10 or more days before SIPO |  | -0.179<br>(1.214) |
| 6 to 9 days before SIPO |  | 1.349<br>(2.305) |
| 1-5 days after SIPO | -1.744<br>(1.291) | -1.133<br>(1.333) |
| 6-10 days after SIPO | -2.527**<br>(1.102) | -1.892*<br>(1.132) |
| 11-15 days after SIPO | -2.741**<br>(1.115) | -2.075*<br>(1.142) |
| 16-20 days after SIPO | -2.633**<br>(1.086) | -1.959*<br>(1.118) |
| 21 or more days after SIPO | -2.830**<br>(1.200) | -2.194*<br>(1.249) |
| Observations | 15,568 | 15,568 |
| R-squared | 0.145 | 0.144 |

Robust standard errors clustered by County-pair × State × Day in parentheses. All economic, demographic, and health controls are included. Models are weighted by county population.
\*\*\* $p < 0.01$ , \*\* $p < 0.05$ , \* $p < 0.1$

**Figure 2.**
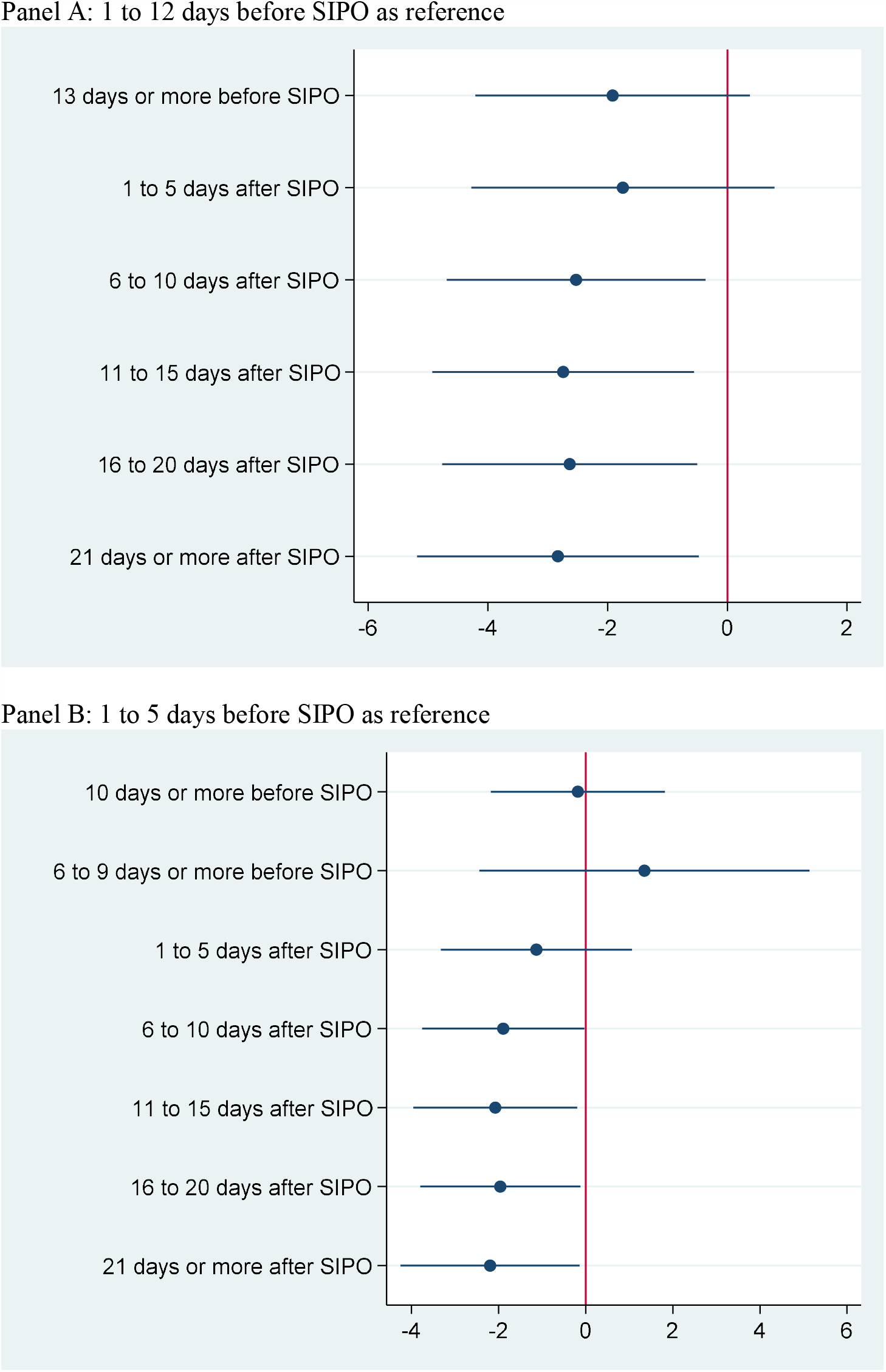
Event study estimates.

## 4. ROBUSTNESS TESTS

### 4.1 Alternate time frame and growth rates

The timeframe for our main analysis is from March 1 to April 25, 2020. Because some of the treated states partially reopened on April 26, 2020, we test the robustness of our main results by relaxing the start date. In Table 5, Models 2, 3, and 4 for start dates of Feb 25^th^, March 15^th,^ and Jan 22^nd,^ 2020, respectively, we find that the SIPO effects are negative and significant and our results are consistent with alternate start dates of the analysis.

**Table 5:**
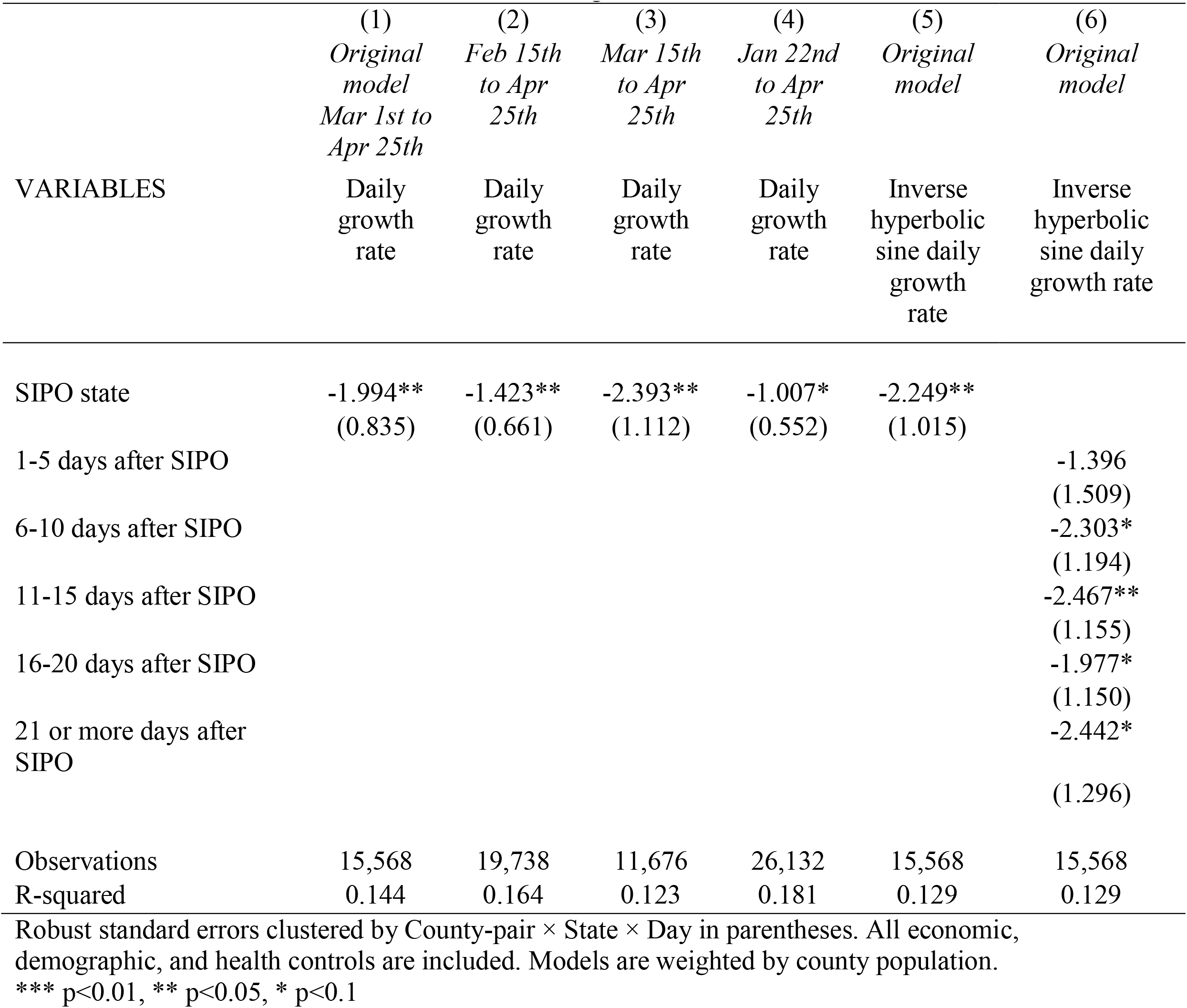
Robustness tests on start dates and alternate growth rate

| VARIABLES | (1)<br>*Original*<br>*model*<br>*Mar 1st to*<br>*Apr 25th* | (2)<br>*Feb 15th*<br>*to Apr*<br>*25th* | (3)<br>*Mar 15th*<br>*to Apr*<br>*25th* | (4)<br>*Jan 22nd*<br>*to Apr*<br>*25th* | (5)<br>*Original*<br>*model* | (6)<br>*Original*<br>*model* |
| --- | --- | --- | --- | --- | --- | --- |
|  | Daily<br>growth<br>rate | Daily<br>growth<br>rate | Daily<br>growth<br>rate | Daily<br>growth<br>rate | Inverse<br>hyperbolic<br>sine daily<br>growth<br>rate | Inverse<br>hyperbolic<br>sine daily<br>growth rate |
| SIPO state | -1.994**<br>(0.835) | -1.423**<br>(0.661) | -2.393**<br>(1.112) | -1.007*<br>(0.552) | -2.249**<br>(1.015) |  |
| 1-5 days after SIPO |  |  |  |  |  | -1.396<br>(1.509) |
| 6-10 days after SIPO |  |  |  |  |  | -2.303*<br>(1.194) |
| 11-15 days after SIPO |  |  |  |  |  | -2.467**<br>(1.155) |
| 16-20 days after SIPO |  |  |  |  |  | -1.977*<br>(1.150) |
| 21 or more days after<br>SIPO |  |  |  |  |  | -2.442*<br>(1.296) |
| Observations | 15,568 | 19,738 | 11,676 | 26,132 | 15,568 | 15,568 |
| R-squared | 0.144 | 0.164 | 0.123 | 0.181 | 0.129 | 0.129 |

Robust standard errors clustered by County-pair × State × Day in parentheses. All economic, demographic, and health controls are included. Models are weighted by county population.
\*\*\* $p < 0.01$ , \*\* $p < 0.05$ , \* $p < 0.1$

**Table 6.** Heterogeneity by economic characteristics

| VARIABLES | (1)<br>Daily<br>growth<br>rate | (2)<br>Daily<br>growth rate | (3)<br>Daily growth<br>rate |
| --- | --- | --- | --- |
| SIPO state | 3.486<br>(8.808) | -4.647**<br>(2.247) | 39.87***<br>(9.050) |
| GINI inequality Index | 14.82<br>(11.05) | 12.16<br>(10.06) | 4.394<br>(10.32) |
| SIPO state × GINI inequality index | -12.37<br>(19.55) | | |
| Unemployment rate | 0.382<br>(0.440) | 0.167<br>(0.442) | 0.414<br>(0.445) |
| SIPO state × Unemployment rate | | 0.678<br>(0.473) | |
| Log of Real GDP | 1.372**<br>(0.534) | 1.252**<br>(0.536) | 2.231***<br>(0.568) |
| SIPO state × Log of Real GDP | | | -2.867***<br>(0.658) |
| Observations | 15,568 | 15,568 | 15,568 |
| R-squared | 0.144 | 0.145 | 0.152 |

Robust standard errors clustered by County-pair × State × Day in parentheses. All controls are included. Models are weighted by county population.
\*\*\* $p < 0.01$ , \*\* $p < 0.05$ , \* $p < 0.1$

We also use an inverse hyperbolic sine transformation of our growth rate outcome and retest our results (Burbidge, Magee, & Robb, 1988; Courtemanche et al., 2020). In Table 5 Models 5 and 6 we find consistent results.

### 4.2 Heterogeneity by economic characteristics

We test whether SIPO had differentially impacted outcomes based on the economic characteristics of the county. We perform heterogeneity tests with income inequality, unemployment rates, and county GDP. We find that SIPO in high-income inequality or unemployment regions did not have any effect on daily growth rates of COVID-19 incidences.

We find that SIPO in higher GDP counties had lower SIPO incidence rates that lower GDP regions. Further research is needed to explore this finding.

### 4.3 Heterogeneity by demographic characteristics

We also test whether SIPO differentially impacted regions by heterogeneity in the race, elderly population, population density (Dave, Friedson, Matsuzawa, Sabia, et al., 2020), and prior social association rates. In Appendix Table B, the inference for the heterogeneity tests is based on plotting the margins, wherein the accounting for direct and interaction effects, the net effect is negligible.

SIPO in counties concentrated with a higher Black/African American, a lower Hispanic population, a higher share of the population who are 65 years and older, or a higher social association rate had a negligible increase in COVID-19 incidence rates. We do not find a statistically different effect of SIPO by higher population density.

### 4.4 Heterogeneity by regional health

It is plausible that a region with poor health infrastructure could be prone to higher incidence rates of COVID-19. We test heterogeneity by prior county primary care physician availability in 2017 (defined as ratio of population to primary care physicians), prior county age-adjusted death rates in 2016 to 2018 (death rate among residents under age 75), and prior county preventable hospitalization rates in 2017 (defined as rate of hospital stays for ambulatory-care sensitive conditions per 100,000 Medicare enrollees). The data for these three variables were obtained from County Health Rankings. In Appendix Table C, our results show that the net interaction effect of these factors was negligible.

Furthermore, controlling for all these three variables (Model 4) in our preferred specification led to similar inferences.

### 4.5 Additional test with Census division

To capture any varying trends of COVID-19 incidence across the regions in the country, we also conduct a robustness test by interacting census division with day fixed effects. Appendix Table D shows that the results were consistent with the main inferences.

## 5. CONCLUSION

In this cross-border county study, we found that the implementation of SIPO was associated with a reduction of 1.99 percent points in the day-to-day growth rate of COVID-19 incidence relative to cross-border county without SIPO. Our estimates imply that the decline is lower than that estimated in previous studies (Courtemanche et al., 2020). The estimates did not vary much by differences in demographic, economic, or health infrastructure conditions. The results are in line with the theoretically expected effects of SIPO on COVID-19 spread, however, the effect sizes are substantially lower than prior studies. Based on Rojas et al. (2020) it possible that proactive closure policies from the school and private sector may lower the effects of SIPO. Our empirical specification only makes it feasible to include the states from southern, mid-west, and mountain regions in the US. While the proposed specification cannot be applied to the western and the north-eastern US where almost all states adopted SIPO, the estimates show marginal benefits of SIPO.

Related to the limitations of the study, measurement bias underscores the count of COVID-19 cases. Variation in availability of testing facilities and daily testing capacities, presence of asymptomatic carriers, general predisposition in the population to get tested, among others, are some of the conditions that may limit a reliable count of daily COVID-19 cases. As such, much of the available data remains right-censored. Though causality is difficult to establish, the border-county pair analysis along with matched-pair county and day fixed-effects and three-way clustering aim to provide additional estimates to the ongoing stream of studies on SIPO and decline of COVID-19. Though closures of offices and schools from across state lines might spillover across border county, such effects would downward bias our estimates and control for matched-pair county fixed-effects control for potential spillovers. In conclusion, we demonstrate that the US COVID-19 pandemic daily growth rate slowed at 1.99 percentage points in SIPO counties relative to non-SIPO counties and suggests smaller benefits of SIPO.

## Data Availability

Data is available with authors upon request

## Online Appendix

**Table A.** List of states and SIPO effective dates used in the analysis

| Name of State | SIPO effective |
| --- | --- |
| Arizona | 3/31/2020 |
| Arkansas | non-SIPO |
| Colorado | 3/26/2020 |
| Idaho | 3/25/2020 |
| Illinois | 3/21/2020 |
| Iowa | non-SIPO |
| Kansas | 3/30/2020 |
| Louisiana | 3/23/2020 |
| Minnesota | 3/27/2020 |
| Mississippi | 4/3/2020 |
| Missouri | 4/6/2020 |
| Montana | 3/28/2020 |
| Nebraska | non-SIPO |
| Nevada | 4/1/2020 |
| North Dakota | non-SIPO |
| Oklahoma* | non-SIPO |
| South Dakota | non-SIPO |
| Tennessee | 3/31/2020 |
| Texas | 4/2/2020 |
| Utah* | non-SIPO |
| Wisconsin | 3/25/2020 |
| Wyoming | non-SIPO |

\* Cities in Sequoyah county in Oklahoma State and Summit County in Utah state implemented SIPO. Those border counties were included in our treated group.

**Table B.** Heterogeneity by regional demographic characteristics

| VARIABLES | (1)<br>Daily<br>growth rate | (2)<br>Daily<br>growth rate | (3)<br>Daily<br>growth rate | (4)<br>Daily<br>growth rate | (5)<br>Daily<br>growth rate |
| --- | --- | --- | --- | --- | --- |
| SIPO state | -2.490***<br>(0.936) | -0.0728<br>(0.968) | -15.15***<br>(3.584) | -1.507*<br>(0.816) | -7.260***<br>(1.707) |
| Share of Black/African American population | 0.0722<br>(0.0758) | 0.124*<br>(0.0668) | 0.154**<br>(0.0675) | 0.137**<br>(0.0689) | 0.157**<br>(0.0676) |
| SIPO state × Share of Blk/Afr Amer pop | 0.0688**<br>(0.0318) |  |  |  |  |
| Share of Hispanic population | 0.120<br>(0.0886) | 0.165*<br>(0.0927) | 0.160*<br>(0.0902) | 0.125<br>(0.0882) | 0.100<br>(0.0881) |
| SIPO state × Share of the Hispanic population |  | -0.170**<br>(0.0835) |  |  |  |
| Share of population age 65 and over | 0.0418<br>(0.151) | 0.0747<br>(0.151) | -0.144<br>(0.157) | 0.0775<br>(0.153) | 0.0997<br>(0.152) |
| SIPO state × Share of pop above 65 |  |  | 0.747***<br>(0.175) |  |  |
| Population density |  |  |  | 0.00162<br>(0.0115) |  |
| SIPO state × Population density |  |  |  | -0.00672<br>(0.00877) |  |
| Social association rate | -0.0311<br>(0.0539) | -0.0652<br>(0.0524) | -0.0757<br>(0.0523) | -0.0546<br>(0.0519) | -0.190***<br>(0.0591) |
| SIPO state × Social association rate |  |  |  |  | 0.446***<br>(0.0927) |
| Observations | 15,568 | 15,568 | 15,568 | 15,568 | 15,568 |
| R-squared | 0.145 | 0.146 | 0.148 | 0.144 | 0.147 |

Robust standard errors clustered by County-pair × State × Day in parentheses. All controls are included. Models are weighted by county population.
\*\*\* p<0.01, \*\* p<0.05, \* p<0.1

**Table C.** Heterogeneity by regional health

| VARIABLES | (1)<br>Daily<br>growth<br>rate | (2)<br>Daily<br>growth<br>rate | (3)<br>Daily<br>growth<br>rate | (4)<br>Daily<br>growth<br>rate |
| --- | --- | --- | --- | --- |
| SIPO state | 2.525*<br>(1.434) | -9.715***<br>(3.027) | -9.547***<br>(2.184) | -2.114**<br>(0.858) |
| Primary care physicians rate | 0.00712<br>(0.0126) |  |  | -0.00764<br>(0.0115) |
| SIPO state × Primary care physician rate | -0.0764***<br>(0.0238) |  |  |  |
| Age-adjusted death rate |  | -0.00374<br>(0.00642) |  | -0.00455<br>(0.00686) |
| SIPO state × Age-adjusted death rate |  | 0.0198***<br>(0.00642) |  |  |
| Preventable hospitalization rate |  |  | -0.000103<br>(0.000192) | 0.000275<br>(0.000189) |
| SIPO state × Preventable hospitalization rate |  |  | 0.00162***<br>(0.000349) |  |
| Observations | 14,168 | 15,232 | 15,456 | 14,112 |
| R-squared | 0.147 | 0.147 | 0.149 | 0.145 |

Robust standard errors clustered by County-pair × State × Day in parentheses. All controls are included. Models are weighted by county population.
\*\*\* p<0.01, \*\* p<0.05, \* p<0.1

**Table D.**
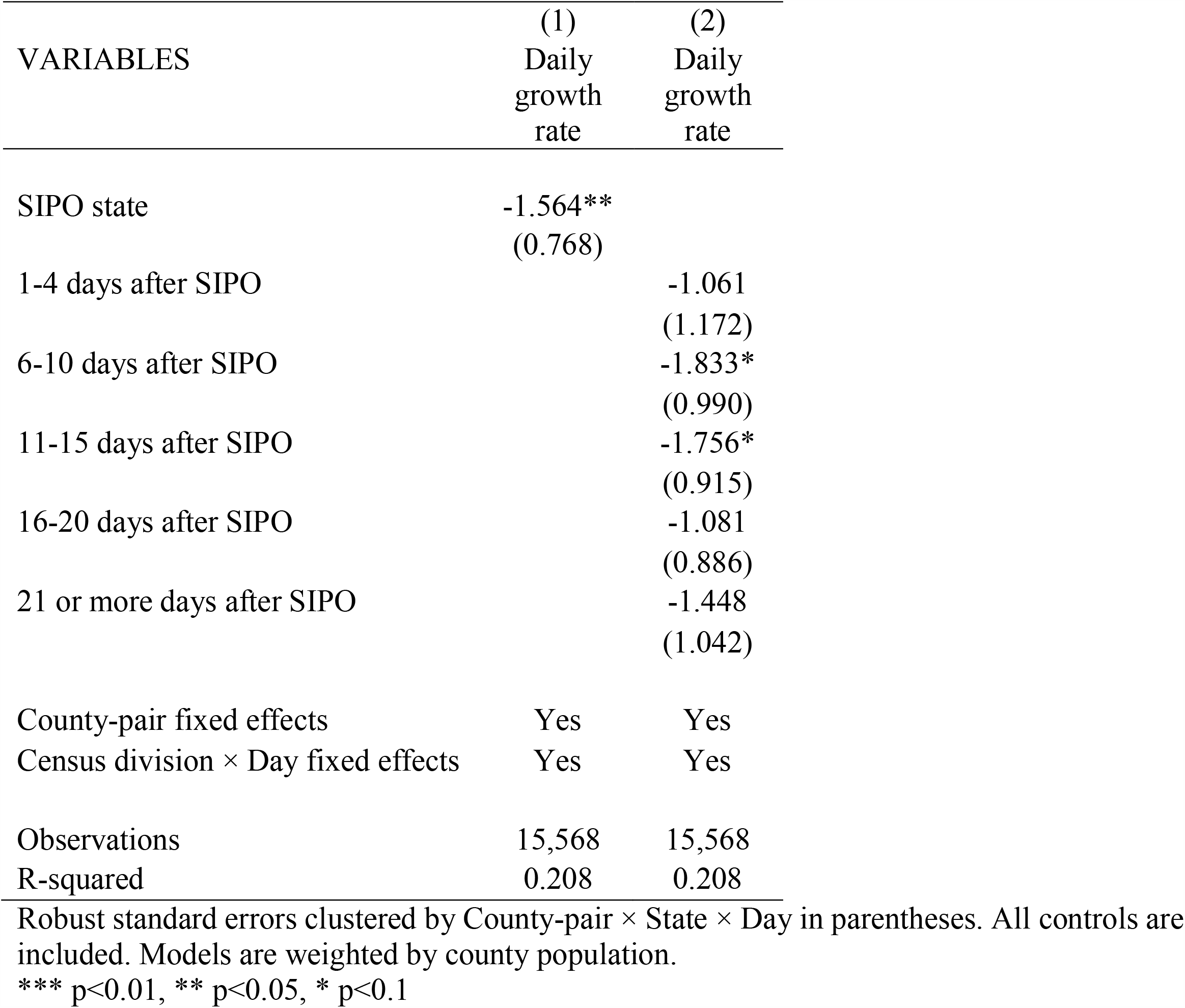
Robustness test with census division × day effects

| VARIABLES | (1)<br>Daily<br>growth<br>rate | (2)<br>Daily<br>growth<br>rate |
| --- | --- | --- |
| SIPO state | -1.564**<br>(0.768) |  |
| 1-4 days after SIPO |  | -1.061<br>(1.172) |
| 6-10 days after SIPO |  | -1.833*<br>(0.990) |
| 11-15 days after SIPO |  | -1.756*<br>(0.915) |
| 16-20 days after SIPO |  | -1.081<br>(0.886) |
| 21 or more days after SIPO |  | -1.448<br>(1.042) |
| County-pair fixed effects | Yes | Yes |
| Census division × Day fixed effects | Yes | Yes |
| Observations | 15,568 | 15,568 |
| R-squared | 0.208 | 0.208 |

Robust standard errors clustered by County-pair × State × Day in parentheses. All controls are included. Models are weighted by county population.
\*\*\* $p < 0.01$ , \*\* $p < 0.05$ , \* $p < 0.1$

## Footnotes

1 County-level incidence data was obtained from https://usafacts.org/visualizations/coronavirus-covid-19-spread-map/

2 SIPO data from https://www.nytimes.com/interactive/2020/us/coronavirus-stay-at-home-order.html

3 The reopening / SIPO expired dates can be obtained from here: https://www.nytimes.com/interactive/2020/us/states-reopen-map-coronavirus.html

4 This includes counties in our sample that borders with multiple counties and sevral counties bordering with a single county were all assigned a unique pair.

